# Capturing SARS-CoV-2 immune landscapes to inform future strategies for COVID-19 vaccination in a high-income setting: a mathematical modelling study

**DOI:** 10.1101/2025.03.18.25324222

**Authors:** Alexandra B Hogan, David J Muscatello, Bette Liu, Gemma Nedjati-Gilani, James G Wood

## Abstract

**Background:** In an era of endemic SARS-CoV-2 transmission, countries are continuing to evaluate how best to schedule ongoing COVID-19 booster vaccinations. Mathematical modelling provides a useful tool to predict the benefit of future vaccination strategies, incorporating the loss of protection due to waning immunity and strain mutation.

**Methods:** We adapted a combined immunological-population transmission model for SARS-CoV-2, to better capture contemporary understanding of exposure- and vaccine-derived immunity, to simulate ongoing endemic transmission of SARS-CoV-2 in a highly exposed high-income setting. We used this model to estimate the impact of targeted booster dose strategies in the older population, both in the context of continued circulation of the current dominant viral strain, and in the presence of a new antigenically distinct variant.

**Results:** We found that at the population level, an annual COVID-19 vaccine booster dose to the 65+ years population at 60% coverage could avert 10–17% of hospitalisations over a single wave, depending on how well-matched the vaccine is to the circulating SARS-CoV-2 strain. With lower coverage of 40%, estimated impact was between 9–12%. A second booster dose to the 75+ population after 6 months was particularly beneficial if a new distinct variant strain increases the magnitude of the wave.

**Conclusions:** This adapted model captures endemic viral transmission and could readily be used to explore vaccine impact across other settings.

## Background

On 5 May 2023, the World Health Organization (WHO) declared that COVID-19 would no longer be considered a public health emergency of international concern (1). The SARS-CoV-2 virus is now characterised by endemic transmission in all countries, with periodic waves due to immunity waning following infection or vaccination, seasonality of infection, and immune escape as the virus evolves (2). Despite the cessation of the emergency, COVID-19 remains an ongoing cause of death globally, with at least 3,000 deaths reported in the 28 days to 10 November 2024 (3). In Australia, COVID-19 continues to cause hospitalisations, intensive care unit (ICU) admissions, deaths, and aged care outbreaks, and was the leading cause of acute respiratory infection mortality across 2022–2024 (4,5). The burden of COVID-19 hospitalisation and severe outcomes remains on average higher than that for influenza (6).

Most of the population globally has likely been exposed to SARS-CoV-2 multiple times over the past four years, and combined with vaccination, this has provided hybrid immunity against infection and disease (7–9). However, protection from both vaccines and infection wanes over time (2). Since the emergence of Omicron, the majority of immune boosting has occurred through exposure to the virus, and vaccination now remains primarily a tool for reducing severe COVID-19 disease and death. This is particularly true in older individuals, those with additional health conditions that increase their risk, and priority populations (10).

In September 2024, WHO’s Strategic Advisory Group on Immunization (SAGE) updated its COVID-19 booster dose advice, recommending 12-monthly booster doses in adults over 50 or 60 years, and 6-monthly boosters in adults over 75 or 80 years, with the age cut-off depending on country, and broader vaccination recommended in individuals with comorbidities and other priority risk groups (11). In Australia, booster doses are now recommended every 12 months for the 65+ years population, and every 6 months for the 75+ years population (12). An additional consideration is vaccine composition; as the virus has evolved, vaccines have been updated to better target the most common circulating strains (13). WHO SAGE recommends being vaccinated with whatever product is available at the time of immunisation. Even though advice on continuing annual or biannual booster doses remains in place, vaccine uptake is moderate. For example, by end of November 2024, the United Kingdom Health Security Agency (UKHSA) reported that around 56% of 65+ year olds had been vaccinated with an autumn 2024 booster dose (14), and according to recent Australian data, around 34% of 65– 74-year olds and around 50% of 75+ year olds had received a COVID-19 booster dose in the last 12 months (15).

Countries now need to plan ongoing scheduling of booster vaccine doses in the context of waning immunity and continual emergence of new variants that evade existing immunity. Mathematical models of COVID-19 transmission and vaccination have been important tools for informing health policy decisions throughout the pandemic (16). However, now that we have moved beyond the emergency response era of COVID-19 control, there is a need to develop models that can inform vaccination planning over longer-term time horizons (17). To do this, models will need to capture the transition to endemic SARS-COV-2 transmission in non-naïve populations with complex and varied vaccination and exposure histories, in the context of high prior infection exposure.

In our study we adapted an existing individual-based population-level transmission model of SARS-CoV-2 transmission and disease, linked to an individual-level immunity model, to capture the relative protection derived from virus exposure and vaccination. Using this model, we generated scenarios for endemic transmission, aiming to capture population-level immunity and levels of infection that would be broadly representative of Australia. We used this model to estimate the future impact of different targeted vaccination scenarios, considering the relative impact of how well-matched the vaccine is to the current circulating variant, the impact of booster dose timing, and the value of vaccination in the context of emergence of new distinct variant strains.

## Methods

### Modelling approach

Our modelling approach is comprised of two steps. First, we use an immunological model that simulates individual-level SARS-CoV-2 immune recognition over time (18). Immune recognition can be boosted by either vaccination or infection and is assumed to decay over time. We then apply a logistic function that captures the relationship between immune recognition and protection from both infection and severe disease (18). Second, to estimate the future benefit of COVID-19 vaccination programs, we simulate these processes at the population level by implementing the individual immunological protection model within an existing individual-based framework of SARS-CoV-2 transmission and disease (19). The modelling framework has been applied previously in the context of estimating the benefit of specific vaccine products, following the emergence of the Omicron variant, but is updated for the present analysis (18,19).

In this present study, we update the combined immunological-population model (19) to capture endemic SARS-CoV-2 transmission in a population with high previous infection incidence and chequered vaccine histories. We do this by reformulating infection- and vaccine-induced protection processes at the individual level, to better represent the differences in protection and durability following exposure versus vaccination, reconstructing the implementation of vaccine-versus infection-derived immunity within the population-level model, and recalibrating the model to capture characteristics of contemporary transmission patterns and recent effectiveness data. The model approach and changes are described in more detail below.

### Immunological model

The dynamics of the immunological model are described in detail elsewhere (18,19) and are summarised as follows. Briefly, we follow the approach presented by Khoury et al (20), in that an individual’s immunity following either infection or vaccination decays exponentially in a biphasic fashion, with a short period of fast decay followed by a longer period of slow decay. This immunity is related to efficacy against infection and severe disease by a logistic function (see Supplementary Material) (20). Expressing the immunological model in this way allows an estimated fold-change reduction in immune recognition (for example by the emergence of a new variant) to be translated to reductions in efficacy against mild and severe disease. In our modelling approach, we conceptualise “immune recognition” to the SARS-CoV-2 virus antigen at the individual level, as a broad level of immunity (which can be comprised of a range of immune mechanisms, including B-cell and T-cell-mediated immunity (21,22)), in response to the currently circulating SARS-CoV-2 variant.

An individual’s level of immune recognition can be boosted by either infection or vaccination. This immune recognition then wanes over time, according to the process described above. For each simulated individual, the infection-acquired and vaccine-acquired levels of immune recognition are allocated separately, and decay independently. The decay dynamics of infection- and vaccine-acquired immune recognition are similar, but we allow for different levels of boosting from infection and vaccination, and additionally account for the extent that a vaccine is matched to the circulating variant (either “well-matched” or “partially-matched”).

At each exposure- or vaccine-induced boost, we assume that the level of immune recognition returns to the maximum level for that type of boost (**Figure 1**). Vaccine- and infection-derived immune recognition are modelled independently, but the level of protection for an individual at each timepoint is assumed to be the maximum of the two. The immune recognition model is fully described in the Supplementary Material.

**Figure 1:**
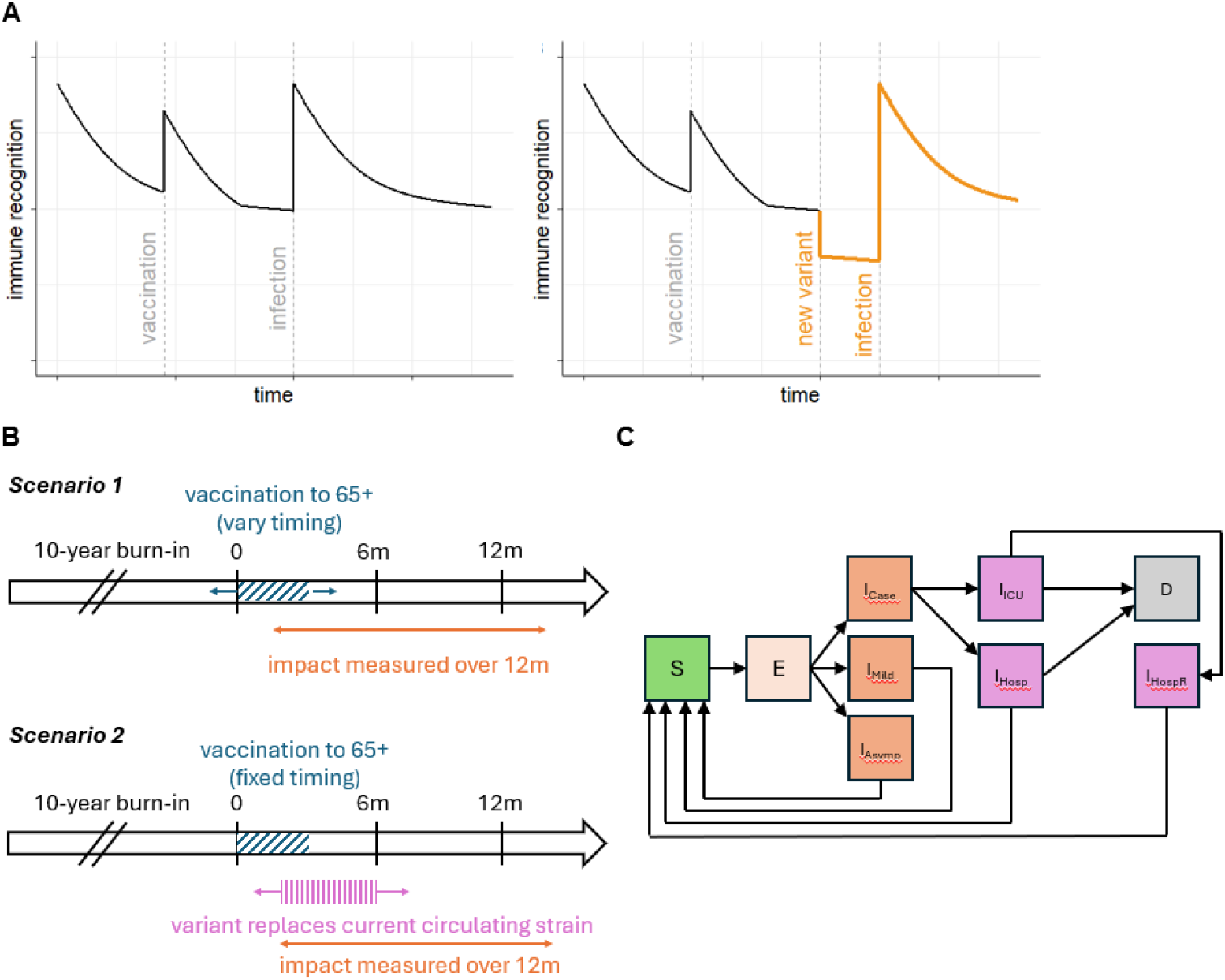
Model schematic and scenarios schematic diagram. (A) Schematic representation of total immune recognition over time at the individual level (formulated as the maximum of vaccine- and infection-induced recognition), following vaccination, and then infection. The right-hand side panel additionally shows the drop in immune recognition following emergence of a distinct variant, illustrated in orange. (B) Schematic representation of the vaccination scenarios implemented using the individual-based population-level model. (C) Schematic flow diagram illustrating the compartmental structure of the population-level SARS-CoV-2 model. Compartments are S (susceptible), E (exposed), I_Case_ (infectious case, prior to hospital admission), I_Mild_ (infectious mild), I_Asymp_ (asymptomatic infection), I_ICU_ (infection requiring ICU care), I_Hosp_ (infection requiring hospital admission), I_HospR_ (recovered following discharge from hospital), and D (death).

### Population-level transmission model

We embedded this individual-level function within a stochastic model of SARS-CoV-2 transmission, severe disease, and vaccination (“safir”), as previously described (19,23). In brief, the model stratifies the population by the following states: susceptible, exposed, infectious (either mild, asymptomatic, or requiring hospitalisation), hospitalised with or without ICU admission, and death (**Figure 1**). Each modelled individual is assigned a 5-year age bin, with bin sizes corresponding to population demography. The total population size is held constant, and births and natural deaths are not simulated. Model parameters are summarised in **Table 1**.

**Table 1:** Summary of fixed SARS-CoV-2 transmission model parameters. Additional epidemiological model parameters are outlined in Table S1.

| Category | Parameter | Description | Value (range tested) | Reference |
| --- | --- | --- | --- | --- |
| Immunological model | $\mu$ | Baseline level of immune recognition<sup>1,2</sup> | 0.22 | (18) |
| | $\delta_I$ | Maximum fold-change increase in immune recognition following infection | 3 (1–5) | Calibrated |
| | $\delta_V$ | Maximum fold-change increase in immune recognition following vaccination | 2 (1–3) | Calibrated |
| | $h_s$ | Half-life of immune recognition decay (short) (days) | 35 | (18) |
| | $h_l$ | Half-life of immune recognition decay (long) (days) | 1000 | Estimated |
| | $t_s$ | Time period for switching (days) | 75 | (18) |
| | $n_i$ | Immune recognition relative to convalescent required to provide 50% protection from mild disease | 0.091 | (18) |
| | $n_s$ | Immune recognition relative to convalescent required to provide 50% protection from hospitalisation | 0.021 | (18) |
| | $k$ | Shape parameter | 2.5 | Estimated |
| | $\sigma_i$ | Standard deviation of individual responses | 0.44 | (20) |
| Setting | $R_t$ | Reproduction number | 3 (3–5) | Estimated |
| | $b_t$ | Transmission coefficient | Calculated from $R_t$ | - |
| | $b_1$ | Amplitude of seasonal forcing | 0.04 (0–0.1) | Calibrated |
| | $c(a, a')$ | Age-structured contact matrix | `squire::get_mixing_matrix (country = "United Kingdom")` | (31) |
|  | - | Demography | From World Population Prospects 2020 data for Australia | (30) |
| Vaccination | $\kappa$ | Maximum coverage within age groups targeted | 40% (20%, 60%) | (14,15) |
|  | - | Rollout period | Uniform over 3 months | - |
|  | - | Age groups targeted (years) | 65+ | (12) |
| Simulation | N | Simulated population size | 1 million | - |
| | $\lambda_{\text{External}}$ | Number of daily importations per million population size | $1 \times 10^{-6}$ | - |
|  | - | Repetitions | 20 | - |
|  | - | Burn-in period | 10 years | - |

<sup>1</sup>calculated as the estimated level of immunity against Delta for the Moderna vaccine third dose of 1.13 from Hogan et al (18), divided by the estimated fold reduction of 5.1
<sup>2</sup>note that all immune recognition level values are expressed as values relative to a theoretical convalescent individual, from Khoury et al (20)

### Model calibration

We previously parameterised the immunological model by fitting to vaccine product-specific effectiveness data from the United Kingdom, in the context of the Delta and Omicron variants (18). However, more recent data highlights some limitations with this approach that we aim to address in this study.

First, the time period of observation following the third vaccine dose in the UKHSA was short, and therefore the decay of vaccine effectiveness following the first booster dose was not well captured (18). Contemporary immunogenicity data suggests a more stable longer-term antibody response following one or more booster doses (24–26). In the present study, we therefore retain the fitted immunological parameters in Hogan et al corresponding to the initial phase of rapid decay (18), and adjust the shape parameter and parameters corresponding to the long period of slow decay to produce a longer half-life.

Second, the initial immunity model was calibrated to a particular category of exposure (i.e. vaccination with a vaccine antigen mismatched to the virus antigen). We assume that the fitted parameters represent efficacy for an ancestral strain-matched vaccine against the Omicron variant (i.e. a significant mismatch) (18). We then introduce a new scaling parameter *δ*, which corresponds to the fold-change increase in immune recognition relative to the baseline fitted parameter set (see **Figure 2**). We define *δ*_*V*_ as the fold-change increase for a vaccine (where the value can be different depending on the level of strain-matching), and *δ*_*I*_ as the fold-change increase for infection, and test values for *δ* between 1 and 5, where *δ*_*I*_ > *δ*_*V*_.

**Figure 2:**
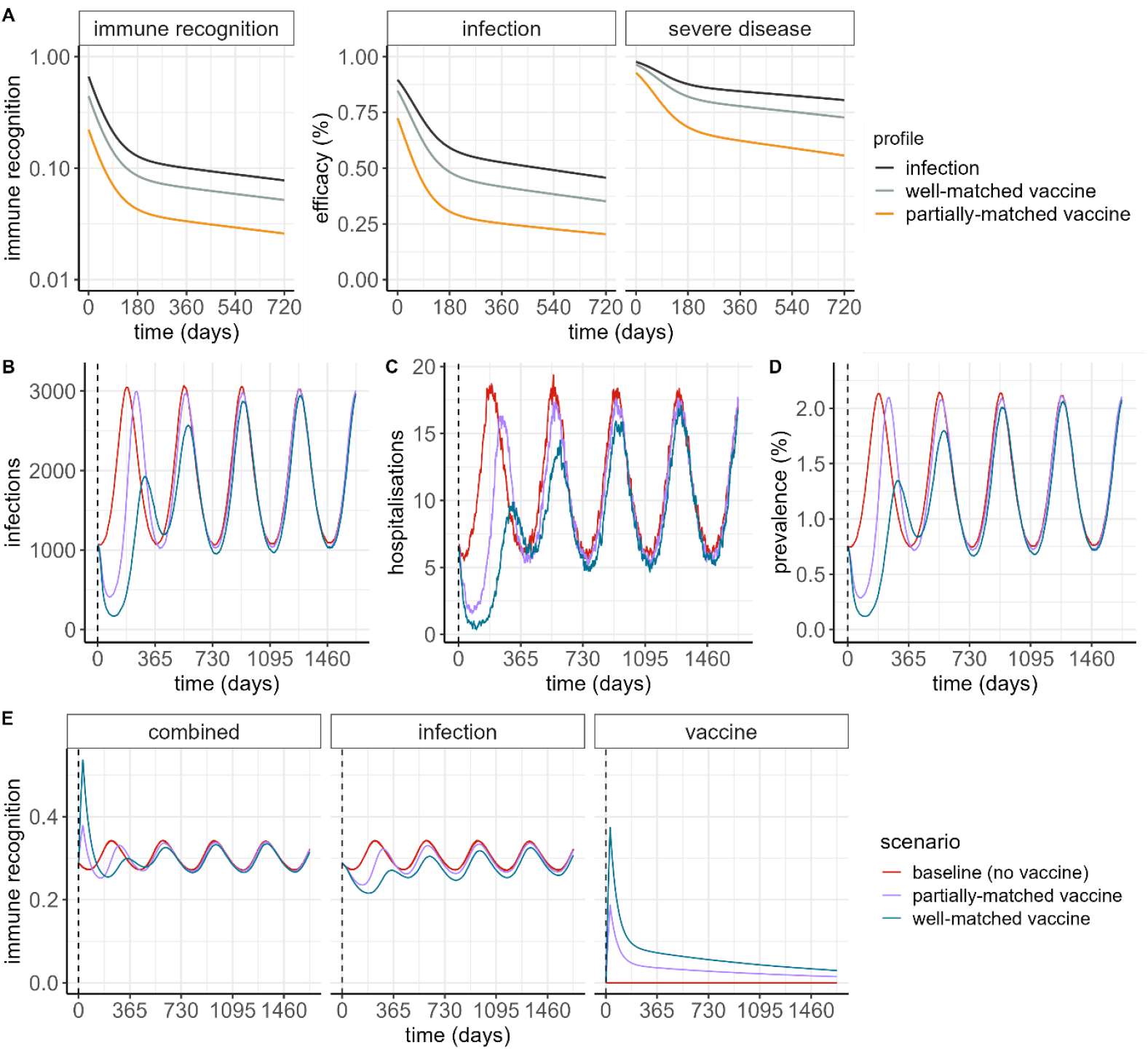
Illustration of population-level impact of individual-level immune recognition and efficacy profiles. (A) Individual-level immune recognition, and corresponding efficacy against infection and severe disease, following infection (black line), and following delivery of a well-matched vaccine (grey line), and a partially-matched vaccine (orange line) at day 0, relative to a naïve individual. (B) Daily infections, and (C) daily hospitalisations, per million population, following a simulation burn-in period of 10 years, and in the absence of any new variant emergence. (D) Prevalence of SARS-CoV-2 infection, corresponding to the epidemic trajectories in panels B and C. (E) Population-level immune recognition. In panels B–E, the vertical dashed line in each panel represents the beginning of the simulated vaccine introduction, where a single vaccine dose is delivered to 80% of the population 15 years and older, over a 2-month period. Note that high vaccination coverage is used in these illustrative simulations to demonstrate the impact of the immune recognition and efficacy mechanism applied in the individual-level transmission model.

Third, the immune profile of the population not receiving a recent booster dose is now markedly different to early 2022, due to repeated exposure to infections. This suggests that protective exposures now are likely to show lower and less durable effectiveness versus comparator cohorts. In order to account for this change in epidemiology, we estimate the magnitude of this fold-change between infection- and vaccine-induced immune protection using vaccine effectiveness data.

For the population-level transmission model, we did not explicitly fit the model to timeseries of reported cases or severe outcomes. With COVID-19 testing and surveillance now limited, notified cases are not representative of underlying community transmission. Additionally, epidemic wave timing has been determined both by waning immunity, and the emergence of new distinct variants (27), which means that the available timeseries are not necessarily reflective of the endemic transmission dynamics the underlying model is aiming to capture. We instead aimed to calibrate the model to contemporary broad epidemiological characteristics.

The calibration process was performed as follows. We first simulate the population-level transmission model in the absence of vaccination, varying the reproduction number *R*_*t*_, the amplitude of seasonal forcing *b*_1_ and the fold-change increase in infection-induced immune recognition *δ*_*I*_. We retain the parameter combination that most closely aligns with the following representative characteristics: (a) an annual population-level attack rate between 0.5 and 1 (representing that on average, individuals are infected every 1 to 2 years), and (b) an annual seasonal peak in hospitalisations that is of a magnitude similar to the recorded annual peak hospitalisations reported at the state level in Australia; (c) endemic transmission throughout the year, with a seasonal peak between two and three times higher compared to the daily infections between epidemics. These characteristics are described in more detail in the Supplementary material, and parameter ranges are outlined in **Tables 1 and S2**. As an additional verification, following calibration of the transmission model, we compared the modelled infection hospitalisation ratio (IHR) by age with IHR estimates from the 2024 UK winter COVID-19 Infection Survey (28).

Second, we implement vaccination in the model to a fixed cohort of individuals, varying the fold-change increase in vaccine-induced immune recognition *δ*_*V*_. We use the model outputs to estimate vaccine effectiveness over a six-month window, and select a *δ*_*V*_ that produces a modelled vaccine effectiveness against infection of approximately 33% over 6 months for a well-matched vaccine (guided by the recent vaccine effectiveness estimate for the Monovalent XBB.1.5 booster of 32.7% (16.1–46.0%) in the UK SIREN healthcare worker cohort (29)).

### Setting and scenarios explored

We fixed the population size as constant and set the relative sizes of the 5-year age groups to correspond to demography for Australia (30). We parameterised age-based contacts using the mean daily number of contacts reported in POLYMOD, a large multi-country population survey, applying the United Kingdom data (31). We simulated transmission for a fixed reproduction number (following the calibration described in the section above) and ran the simulation for a burn-in period of 10 years, to reach endemic equilibrium in the absence of emergence of novel variants, before simulating the scenarios outlined below.

We first explored the impact of routine COVID-19 vaccination, assuming no new antigenically distinct variant emergence. To do this, we simulated the rollout of a COVID-19 vaccine booster dose over a 3-month period, varying the level of coverage, and estimated the impact in terms of averted infections and hospitalisations over a 12-month timeframe. We explored two vaccine delivery programs: either a single dose delivered to the 65+ years population, or a single 65+ years dose with an additional booster dose to the 75+ years population after 6 months. We also explored the impact of varying vaccination timing (either before or after the start of an annual wave of infection), and of the degree of vaccine matching to the circulating variant.

Second, we explored the impact of routine COVID-19 vaccination, in the context of emergence of a new distinct variant of concern (where the magnitude of immune escape is similar to that from Delta to Omicron), allowing the timing of variant emergence to vary relative to vaccination. These scenarios are summarised in **Figure 1B**.

## Data availability

The R package used to undertake the COVID-19 model simulations is available with a description at https://github.com/abhogan/safir_immunity. All code to replicate the analysis and figures in this paper is available at https://github.com/abhogan/covid_endemic_vaccine.

## Results

### Model calibration

We found that for a reproduction number of *R*_*t*_ = 3 and a fold change in immune recognition from infection of *δ*_*I*_ = 3, combined with an amplitude of seasonal forcing of *b*_1_ = 0.04, the modelled annual attack rate was approximately 0.7 (corresponding to approximately two-thirds of the population being infected each year), with an annual hospitalisation epidemic peak corresponding to around 20 per million per day, in line with our calibration criteria (**Figure S3**). Increasing both the transmission level and immunity following infection simultaneously (i.e. *R*_*t*_ and *δ*_*I*_) produced similar attack rates, therefore we retained the parameter combination of *R*_*t*_ = 3 and *δ*_*I*_ = 3 for the main simulations. The model produced annual epidemic waves of infection, and an average annual prevalence of approximately 1.4% (**Figure 2B–D**). We found that the modelled IHR aligned reasonably well with estimates of IHR from the UK COVID-19 Infection Study (**Figure S4**) (28).

We estimated that *δ*_*V*_ = 1 corresponded to vaccine effectiveness against infection over six months of approximately 21% (denoted “unmatched vaccine”), and *δ*_*V*_ = 2 corresponded to vaccine effectiveness over six months of approximately 36% (denoted “well-matched vaccine”). We categorised *δ*_*V*_ = 1.5 as corresponding to a “partially-matched vaccine”. These immune recognition and efficacy profiles are plotted in **Figure 2A**. Illustrative population-level vaccine impact for the well-matched and partially-matched vaccine profiles, with vaccination delivered to ages 20+ years at 80% coverage are shown in **Figure 2B–D**, with the corresponding population-level immune recognition over time plotted in **Figure 2E**. Fixed and calibrated parameters are summarised in **Tables 1 and 2**.

### Impact of routine vaccination

We estimated that overall, a single vaccine dose delivered to the 65+ population at 40% coverage could avert around 8%–12% of COVID-19 hospitalisations over a single seasonal epidemic wave (corresponding to 32–48 averted per 100,000 individuals), depending on the degree that the vaccine is matched to the dominant circulating strain (**Figure 3, Figure S5**). Up to 10%–17% of hospitalisations may be averted if a higher level of vaccine coverage of 60% is achieved (around 40–70 averted per 100,000 individuals) (**Table S3**). An additional vaccine dose delivered to the 75+ population was estimated to be beneficial (averting an estimated 10 additional hospitalisations per 100,000 individuals at 60% coverage). However, for the scenarios modelled, a two-dose strategy with a partially-matched vaccine averted more hospitalisations overall compared to a well-matched vaccine delivered to only the 65+ population (**Table S3**).

**Figure 3:**
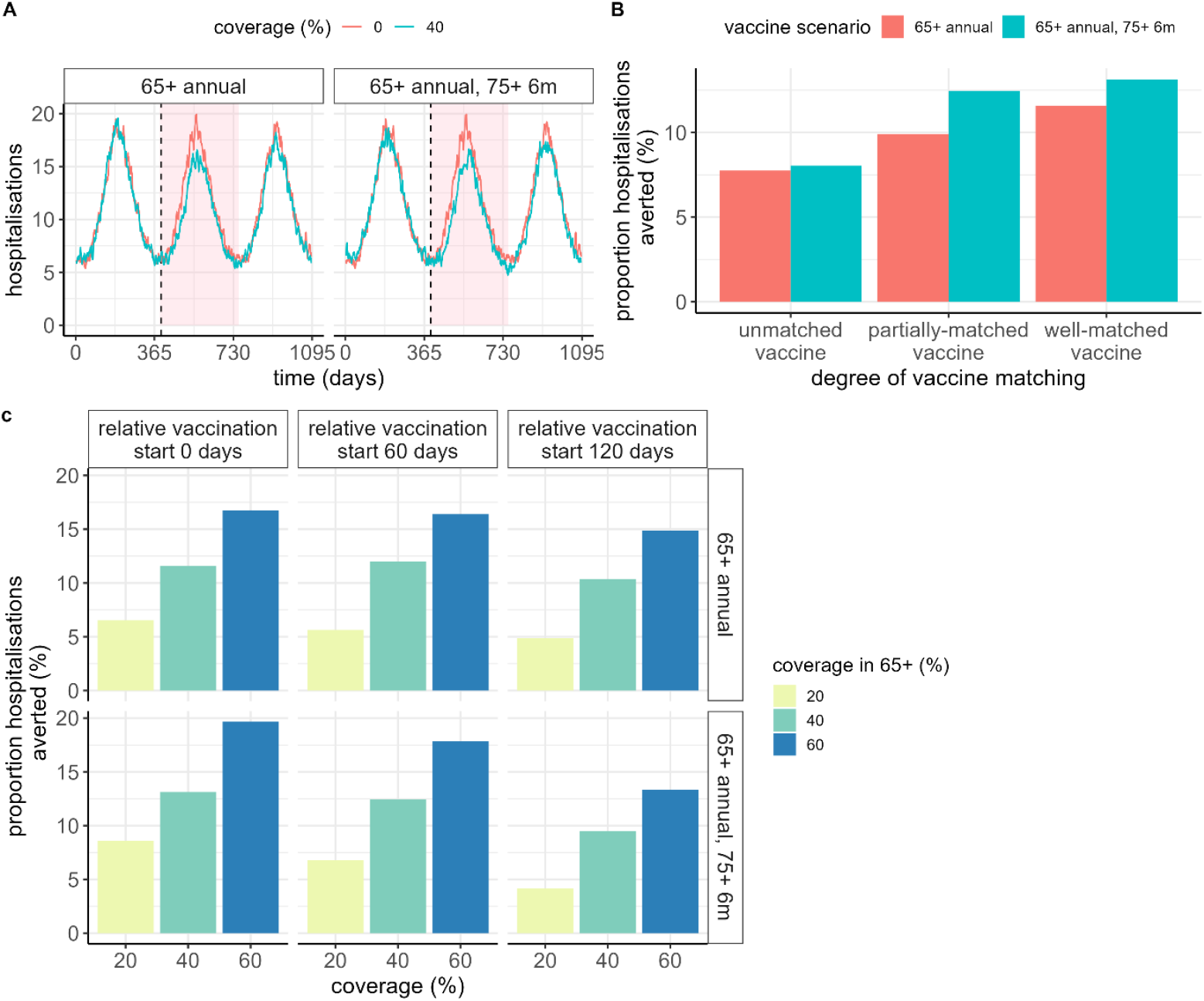
Impact of routine vaccination over a 12-month period. (A) Daily hospitalisations per million population for a scenario with no vaccination (red line), compared to a vaccine delivered at 40% coverage (blue line), where the start of vaccination delivery is shown with a vertical dashed line, and the pink shaded region denotes the time window over which events are aggregated to quantify impact. (B) Proportion of hospitalisations averted for three vaccine product scenarios: either an unmatched vaccine, with *δ*_*V*_ = 1, a partially-matched vaccine, with *δ*_*V*_ = 1.5, or a well-matched vaccine, with *δ*_*V*_ = 2. (C) Proportion of hospitalisations averted for coverage levels of 20%, 40% and 60%, assuming delivery of a well-matched vaccine, for different timings of vaccination commencement relative to the trough in seasonal infections, where the “0 days” scenario is reflected in panel A. In all panels, “65+ annual” refers to a single vaccine dose delivered to the 65+ years population, and “65+ annual, 75+ 6m” represents the same scenario but with an additional dose delivered to the 75+ years population after a six-month period. Additional trajectories as in panel A, but with vaccination commencing at different timepoints relative to the epidemic wave, are shown in **Figure S5**. Values are shown in **Table S3**.

We found that timing of vaccine implementation relative to a seasonal wave is important. Assuming vaccination is rolled out uniformly over a 3-month period, the maximum benefit was achieved if vaccination is commenced around 6 months prior to the estimated epidemic peak, with only around two-thirds of the benefit achieved if vaccination commences 4 months later, or well into the epidemic wave (**Figure 3C, Table S3**).

At the individual level, we estimated that receiving a partially matched vaccine earlier in the season was relatively comparable to a later well-matched vaccine delivered around the peak of the season, in terms of risk of hospitalisation within that epidemic season **(Figure S6**). However, a well-matched vaccine provided more long-lasting benefit into the subsequent COVID-19 epidemic wave.

### Impact of vaccination in context of new variant emergence

We found that even a poorly matched vaccine delivered to 60% of the eligible population provides considerable population-level benefit if a new variant emerges (**Figure 4, Figure S7, and Figure S8)**. If a second vaccine dose is delivered to the 75+ population 6 months after the first dose, at the same coverage, close to 20% of hospitalisations could be averted. Although the total population-level impact was estimated to be higher if the second dose is updated to match the new variant strain, an additional dose to the highest risk group with the same vaccine could potentially avert up to an additional 10% of hospitalisations compared to a single-dose policy (**Table S4**).

**Figure 4:**
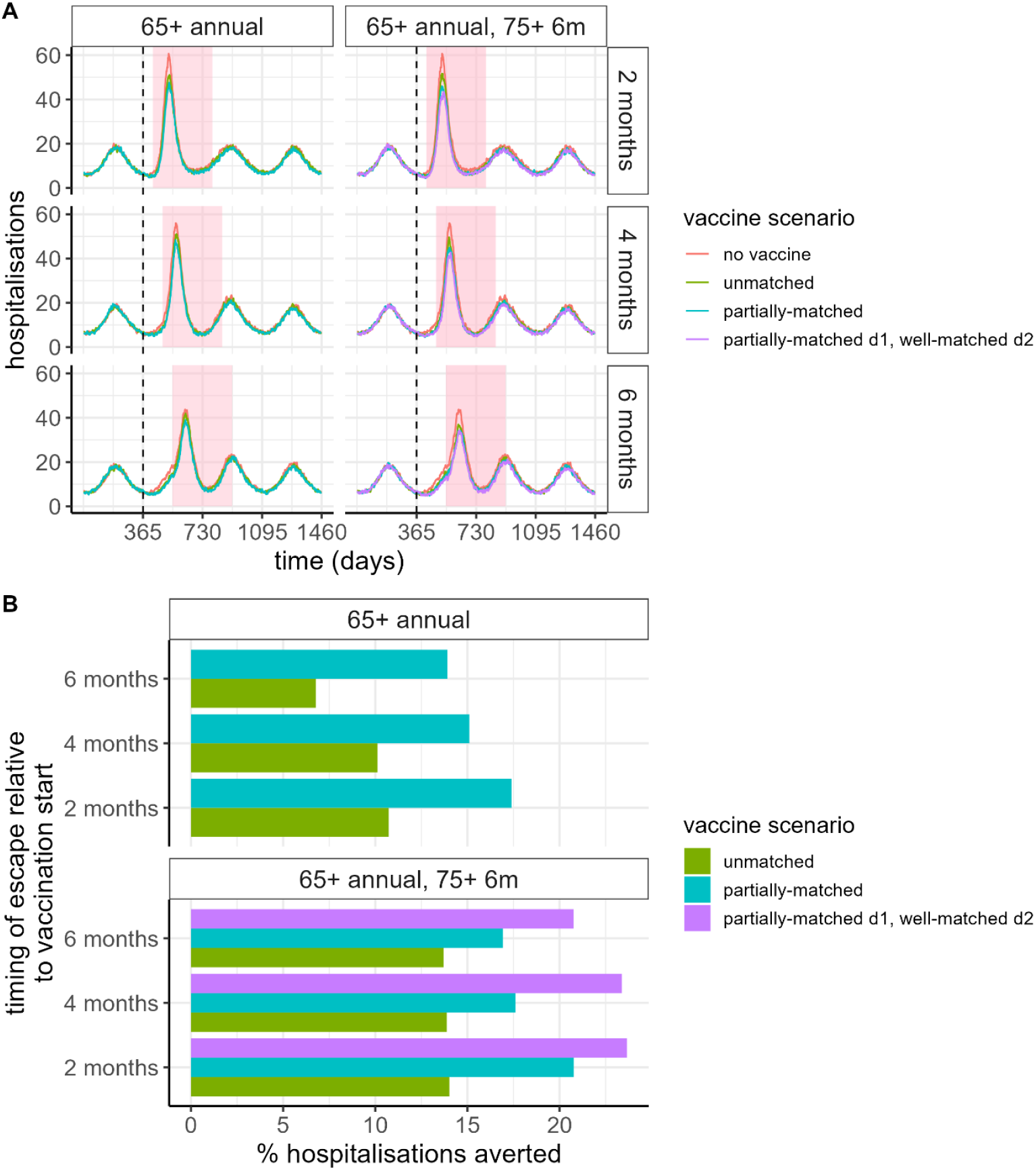
Impact of vaccination in the context of emergence of a new distinct variant strain. (A) Daily hospitalisations, following variant emergence at either 2, 4, or 6 months following vaccination (where the start of vaccine delivery is shown with a vertical dashed line). (B) Vaccine impact in terms of percentage of hospitalisations averted, where the total impact for each timing scenario is aggregated over a 12-month period following variant emergence (shown as the pink shaded regions in panel A). Coloured lines and filled bars represent four scenarios: no vaccine (red); an unmatched vaccine (green), with *δ* = 1.5, a partially-matched vaccine, with *δ* = 2 (teal), and a partially matched first dose to the 65+ years population, with a second dose after 6 months delivered to the 75+ years population with a vaccine dose that is well-matched to the new variant strain (purple). Coverage is assumed to be 60%. Values are shown in **Table S4**.

## Discussion

To date, mathematical models of SARS-CoV-2 transmission and vaccination have largely been formulated in the context of pandemic COVID-19, whereas the present need is to inform longer-term COVID-19 vaccination policies. In our study, we refined a model of individual-level immunity to incorporate the most recent evidence on the effectiveness of vaccines and hybrid immunity. Using this model, we captured contemporary endemic patterns of SARS-CoV-2 infections and hospitalisations observed in Australia and quantified the benefits from variations of an ongoing COVID-19 booster program in older adults. We demonstrated that vaccine coverage and the degree that a vaccine is matched to the current circulating strain are key determinants of COVID-19 vaccine benefit. While optimising the timing of a booster dose relative to the seasonal wave can modify impact, receipt of a vaccine at any time during the season is beneficial.

In Australia, booster doses are now recommended every 12 months for the 65+ years population, and every 6 months for the 75+ years population (12). Uptake has generally declined over time; around 43% of individuals aged 75+ years received a COVID-19 booster dose over the 12 months to January 2025, and in individuals 18+ years, around half as many booster doses were delivered in 2024 compared to 2023 (32–34). We found that a partially matched vaccine delivered once per year to the 65+ years age group at 40% coverage likely averts about 10% of COVID-19 hospitalisations across the population, relative to a no-vaccination scenario. An additional booster to the 75+ population after 6 months can deliver targeted protection to the age group at highest risk of severe disease, and with higher coverage, a well-matched vaccine can avert around 20% of hospitalisations relative to a scenario without vaccination.

In our study we examined the importance of both timing and coverage, and found that for realistic values of these parameters, achieving higher coverage with a booster dose program was more influential in averting hospitalisations compared to the timing of vaccine delivery in advance of the COVID-19 season. In Australia, doses are typically delivered throughout the year, with eligibility based on time since last booster, and although uptake tends to increase with SARS-CoV-2 activity (32), attempting to control timing through policy may be difficult.

Instead, our study indicates that strategies that focus on increasing booster dose uptake in older Australians may be both more effective and efficient in reducing the COVID-19 disease burden. With novel combination vaccines now in the development pipeline (35,36), enabling concurrent protection against multiple respiratory viruses, there may be future opportunities to efficiently increase COVID-19 vaccine booster dose coverage.

While SARS-CoV-2 evolution is unpredictable, even the emergence of novel antigenic variants such as JN.1 have not led to disease burdens comparable to waves between 2020–22, likely because of high existing individual and population-level immunity, particularly to disease consequences of infection (37). However, the emergence of an antigenically distinct variant still has the potential to cause an unanticipated, rapid epidemic wave. In our study, we found that in the event of such an occurrence, vaccine impact is substantially increased, even if a new variant emerges 6 months after vaccine delivery, noting that the benefit of COVID-19 booster doses would vary depending on the characteristics of a novel variant.

Our study has several limitations. First, in terms of calibration, we elected to take a qualitative approach in requiring the model to mimic observed trends in seasonal peaks in hospitalised cases, and the estimated annual incidence rate. This was chosen instead of calibration to case time series data because of uncertain temporal changes in case definitions and testing practices over the period of interest, along with impacts of novel variants, which the model excludes. Second, while the longer-term rate of decay in the previously fitted immunity model was not well-characterised, the calibration source has not been updated with sufficient regularity to facilitate refitting. We therefore revised our immunity model calibration approach and used multiple data sources to validate the underlying immunity profile for protection following exposure, in order to represent a more durable long-term immune response. Other profiles of longer-term immunity may also be plausible. Third, by design our calibrated model produces a single annual (winter) peak, whereas in some settings, including in Australia, an additional smaller (summer) wave has previously been observed. However, given this is based on only two years of observations, and out-of-season waves have been at least partially driven by the emergence of new distinct variants, stability of this pattern is not yet established.

Now that we have moved beyond pandemic-era COVID-19, it is timely for the benefit of vaccine programs to be assessed, as they are integrated into routine national immunisation programs. This model of SARS-CoV-2 transmission and vaccination more accurately captures recent data on protection derived from infection and vaccination and reproduces temporal patterns of the magnitude currently observed in Australia. As demonstrated here, the model can be used to quantify the benefits of ongoing COVID-19 vaccination programs in priority and higher risk groups. This model and calibration approach could be readily applied to other settings, by varying the transmission, seasonality, and demography parameters. There is a need for future work to consider alternative temporal dynamics – it is possible that other epidemic patterns may emerge, this may differ between climatic regions. Finally, with combined vaccines that simultaneously target multiple respiratory pathogens now in development (35,36), dynamic models that capture individual and population immunity across multiple viruses will be required to help determine the most efficient and effective vaccine programs to reduce respiratory disease burden.

## Data Availability

https://github.com/abhogan/safir_immunity

## List of abbreviations

COVID-19: Coronavirus disease
SARS-CoV-2: severe acute respiratory syndrome coronavirus 2
WHO: World Health Organization
UK: United Kingdom
UKHSA: United Kingdom Health Security Agency
WHO SAGE: World Health Organization Strategic Advisory Group on Immunization
VFR: variant fold reduction
AR: attack rate.

## Authorship statement

Designed the study: ABH, JGW; Code development: ABH, GNG; Performed the analysis: ABH; Interpreted the results: ABH, JGW, DM, BL; Drafted the manuscript: ABH; Contributed to manuscript and approved final version: all authors.

## Acknowledgements

This research includes computations using the computational cluster Katana supported by Research Technology Services at UNSW Sydney.

## Supplementary material

### S1. Additional methods

#### Immunological model

An individual’s level of immunity is comprised of infection-induced and vaccine-induced immune recognition. Each of these types of immunity are tracked independently at the individual level, and can be boosted to different thresholds, but follow the same dynamics of decay over time. This model is derived from Khoury et al (20), and is described in the context of the population-level model in Hogan et al (18,19).

Infection-induced and vaccine-induced immune recognition (denoted by *I* and *V* subscripts respectively) are estimated as part of the model calibration process, and the maximum values of these types of immune recognition are defined as

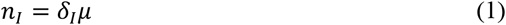

and

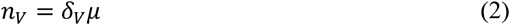

where the parameter *μ* represents the immune level estimated in (18), here referred to as the “baseline” level of immune recognition (see Table 1), and *δ*_*I*_ and *δ*_*V*_ correspond to the maximum fold-change increase in immune recognition following infection and vaccination respectively. The values of *δ*_*I*_ and *δ*_*V*_ are estimated during model calibration.

Each category of immune recognition decays over time according to the biphasic exponential functions

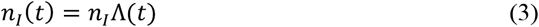

and

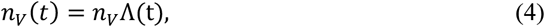

where

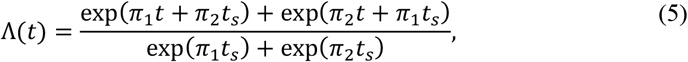

and where *n*_*x*_ is the initial level of immune recognition for either infection or a vaccine, *π*_1_ = −ln(2)/*h*_*s*_ is the rate for the initial period of fast antibody decay; π_2_ = −ln(2)/*h*_*l*_ is the rate for the period of slow decay; *t* is the time since the last infection or vaccine dose; and *t*_*s*_ is the period of switching between the fast and slow decays.

Upon either recovery from infection, or following vaccination, an individual’s immune recognition returns to the maximum level (i.e. the value of Λ(*t*) is reset to 1 for either infection or vaccination), and the decay process recommences. Individual-level variation is accounted for in that each time either an infection or vaccine event occurs, the value of *n*_*I*_ or *n*_*V*_ is drawn from a Normal distribution.

Upon emergence of a new distinct variant strain, immune recognition is reduced linearly over a two-month window, such that after this time window, immune recognition is reduced by a scaling factor termed the “variant fold reduction”, or VFR (initially described in Hogan et al (19)), where

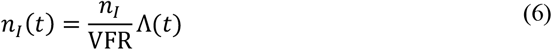

and

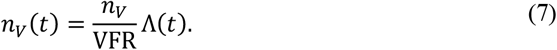

For a vaccine dose that is matched to the new variant strain, the VFR is divided by a scaling factor *ψ* to account for the additional effectiveness of an adapted vaccine. Following infection with a new variant strain, the level of infection-induced immune recognition returns to the maximum level without VFR scaling (i.e. to represent well-matched infection-induced immunity against the current circulating strain).

At each timestep, the total level of immune recognition *n*_*T*_ is calculated as the maximum of the infection- and vaccine-induced levels, such that

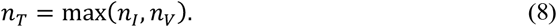

We then assume a logistic relationship between the total level of immune recognition *n*_*T*_ and effectiveness *ϵ*_*m*_, such that

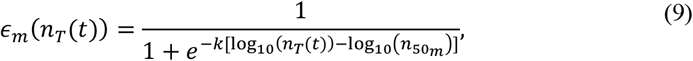

where *k* is the shape parameter and *n*_50*m*_ is the immune recognition required to provide 50% protection against each endpoint (infection (*m* = 1) and hospitalisation (*m* = 2)).

The parameters and their values are described in Table 1.

#### Transmission model

We adapted an existing individual-based population model of SARS-CoV-2 transmission and vaccination (“safir”), to capture the modified immunological model (19). The compartmental model structure is identical to that previously described, but we provide a summary of the model below.

The model stratifies the population into the following epidemiological states (19):

- *S* = uninfected
- *E* = exposed to infection but not yet infectious
- *I*_Mild_ = infected and infectious with mild symptomatic infection that does not require hospitalisation
- *I*_Asymp_ = infected and infectious with asymptomatic infection
- *I*_Case_ = infected and infectious with disease that will require hospitalisation
- *I*_Hosp_ = cases that have been hospitalised in a general ward bed
- *I*_ICU_ = cases that have been admitted to an intensive care unit (ICU)
- *I*_HospR_ = cases that have been stepped down from ICU into a general ward bed for recovery
- *D* = cases that have died.

In the transmission model, infection is initiated into the population at the start of the simulation (with a seed of 10 infections). The model is run with a discrete time step of 0.25 days across a simulation size of 1 million agents. We additionally include a constant external force of infection, representing external infections from outside the modelled (closed) population. All state durations are modelled using an Erlang distribution with shape parameter 2. Transition rates between states are summarised in Table S1, with parameter definitions in Table S2.

**Table S1:** Transmission model state transitions. ICU: intensive care unit. Table is reproduced from (19). Parameters are defined in Table S2.

| Event | State transition | Transition rate |
| --- | --- | --- |
| Infection | $S \rightarrow E$ | $\lambda_{\text{External}} + \lambda(a, n_T, t)$ |
| Latent to asymptomatic infection | $E \rightarrow I_{\text{Asymp}}$ | $\phi_0(a)(1 - \phi_1(a))\alpha$ |
| Latent to mild symptomatic infection | $E \rightarrow I_{\text{Mild}}$ | $(1 - \phi_0(a))(1 - \phi_1(a))\alpha$ |
| Latent to severe infection | $E \rightarrow I_{\text{Case}}$ | $\phi_1(a)\alpha$ |
| Severe case to ICU | $I_{\text{Case}} \rightarrow I_{\text{ICU}}$ | $\phi_2(a)\gamma_2$ |
| Severe case to hospital | $I_{\text{Case}} \rightarrow I_{\text{Hosp}}$ | $(1 - \phi_2(a))\gamma_2$ |
| ICU to death | $I_{\text{ICU}} \rightarrow D$ | $\mu_2(a)\gamma_{4,0}$ |
| Hospitalised to death | $I_{\text{Hosp}} \rightarrow D$ | $\mu_1(a)\gamma_3$ |
| ICU to recovered general ward bed | $I_{\text{ICU}} \rightarrow I_{\text{HospR}}$ | $(1 - \mu_2(a))\gamma_{4,1}$ |
| ICU recovered to susceptible | $I_{\text{HospR}} \rightarrow S$ | $\gamma_5$ |
| Hospitalised to susceptible | $I_{\text{Hosp}} \rightarrow S$ | $(1 - \mu_1(a))\gamma_3$ |
| Symptomatic case to susceptible | $I_{\text{Mild}} \rightarrow S$ | $\gamma_1$ |
| Asymptomatic case to susceptible | $I_{\text{Asymp}} \rightarrow S$ | $\gamma_1$ |

We assume that only infections in the community contribute to onward infection (i.e. the states *I*_Asymp_, *I*_Case_, and *I*_Mild_) and that social mixing is governed by an age-structured contact matrix *c*(*a, a*^′^). The total force of infection acting on each individual is given by

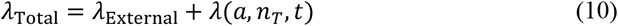

where

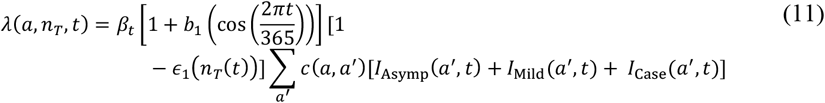

and *β*_*t*_ represents the transmission coefficient, *b*_1_ represents the amplitude of seasonal forcing, *ϵ*_1_ represents efficacy against infection, and *n*_*T*_(*t*) represents the total level of immune recognition.

#### Model calibration

As outlined in the main text, we aimed to calibrate the model so as to reproduce characteristics relating to (a) population-level attack rate; (b) the magnitude of epidemic hospitalisations peaks; and (c) endemic infections, using available Australian surveillance data. These three calibration characteristics are described in more detail as follows.

##### Annual population-level attack rate

No Australian times-series data on incidence or prevalence of infection was available to calibrate cumulative or incident infections. While serosurveys were conducted across 2022 (38), these were not continued into the more relevant post-Omicron era of 2023 and 2024. As such, we relied on data from the UK Covid-19 Infection Survey and estimated that about 25% of the population were infected over the 2023–24 winter wave (a wave that was comparable in size to two previous waves in the same 12 month period) suggesting that based on longer-term sustained levels of SARS-CoV-2 infection in the UK, an annual attack rate of between 0.5–1 (i.e. individuals infected between every one and two years) is appropriate (39).

##### Seasonal hospitalisations

We obtained, where possible, time series hospitalisations data from Australia by state over the years 2023–24. Data from Queensland Health on the number of persons hospitalised with COVID-19 indicates a 2024 winter peak in hospitalisation occupancy of approximately 350-400 persons (40). Assuming a Queensland population of approximately 5.2 million and an average hospital stay of 5 days, this translates to a seasonal peak of around 15 hospital admissions per million per day. Using similar data from Government of Western Australia Department of Health (where data points to a maximum hospital occupancy of approximately 200–250 persons) (41), a state with a population of approximately 2.5 million people, the data translates to a seasonal peak of around 20 hospital admissions per million per day. Finally, data from Victoria, a state with a population of 7 million, indicates a peak seasonal COVID-19 hospital occupancy of about 350-450 (42), translating to around 15 hospital admissions per day. We therefore aimed to reproduce maximum daily hospitalisation incidence of 15 to 20 per million population.

##### Infection characteristics

SARS-CoV-2 infection remains very common, and while seasonal epidemics occur, transmission of the virus is now generally sustained throughout the year (see, for example, UK respiratory virus surveillance data, indicating a baseline level of SARS-CoV-2 positive tests being maintained throughout the year (14), and positive laboratory confirmation data from the Australian National Notifiable Diseases Surveillance System, shown in **Figure S1** (6)).

#### Vaccine allocation

Age-based vaccine allocation is implemented in the transmission model, where the user specifies the target population (here the 65–69, 70–74, 75–79 and 80+ years groups), the day that vaccination commences, the rollout period for a single dose (here 90 days), and the time window between vaccine doses (here 180 days).We then use this information to calculate the number of vaccine doses to be distributed each day over the rollout window, so that doses are administered at a constant rate. Age groups are vaccinated sequentially starting with the oldest age group.

**Figure S1:**
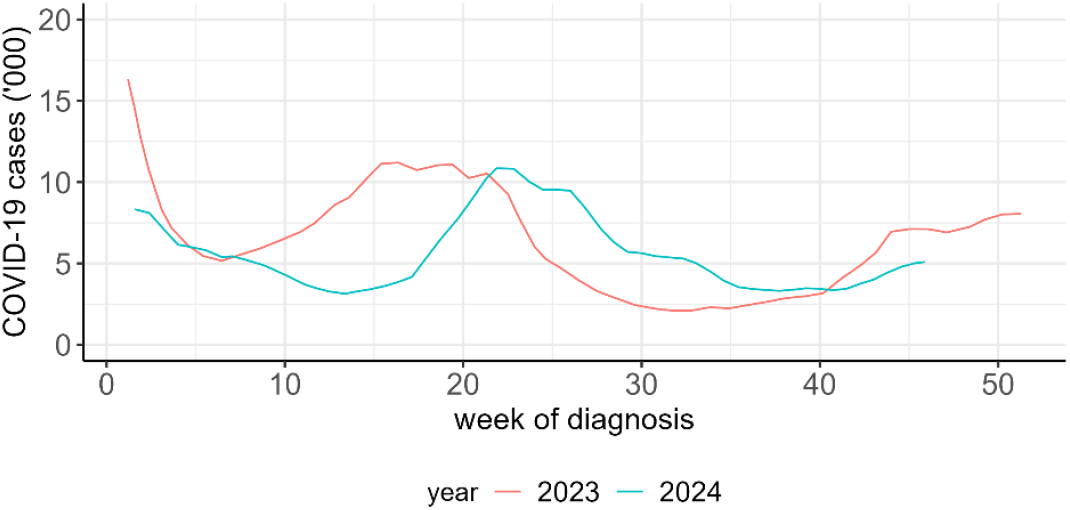
Number of laboratory-confirmed COVID-19 cases notified to the Australian National Notifiable Diseases Surveillance System, by year and week of diagnosis, from 1 January 2023 to 17 November 2024. Data extracted from the Australian Respiratory Surveillance Report 16 – 21 October 2024 to 17 November 2024 (6), using WebPlotDigitizer software (43).

**Table S2:** Additional epidemiological model parameters.

| Parameter | Description | Value (range tested) | Reference |
| --- | --- | --- | --- |
| $1/\alpha$ | Mean latent period | 4.6 days | (44,45) |
| $1/\gamma_1$ | Mean duration of mild or asymptomatic infection | 7 days | (45) |
| $1/\gamma_2$ | Mean duration of severe infection prior to hospitalisation | 7 days | (45) |
| $1/\gamma_3$ | Mean duration of hospitalisation for non-critical cases if survive | 9 days | Assumption |
| $1/\gamma_{4,1}$ | Mean duration in ICU if survive | 14.8 days | (44) |
| $1/\gamma_{4,0}$ | Mean duration in ICU if die | 11.1 days | (44) |
| $1/\gamma_5$ | Mean duration in recovery after ICU | 3 days | Assumption |
| $\mu_1(a)$ | IFR in the absence of immunity (i.e. for a naïve individual) for non-ICU | Age-dependent | Table S2 in Hogan et al 2021 (44) |
| $\mu_2(a)$ | IFR in the absence of immunity (i.e. for a naïve individual) for ICU | Age-dependent | Table S2 in Hogan et al 2021 (44) |
| $\phi_0(a)$ | Proportion of mild infections that are asymptomatic | 0.2 | Estimated |
| $\phi_1(a)$ | Probability of developing severe disease requiring hospitalisation in the absence of immunity | Age-dependent | Table S2 in Hogan et al 2021 (44) |
| $\phi_2(a)$ | Proportion of hospitalisations requiring ICU | Age-dependent | Table S2 in Hogan et al 2021 (44) |

### S2. Additional results

**Figure S2:**
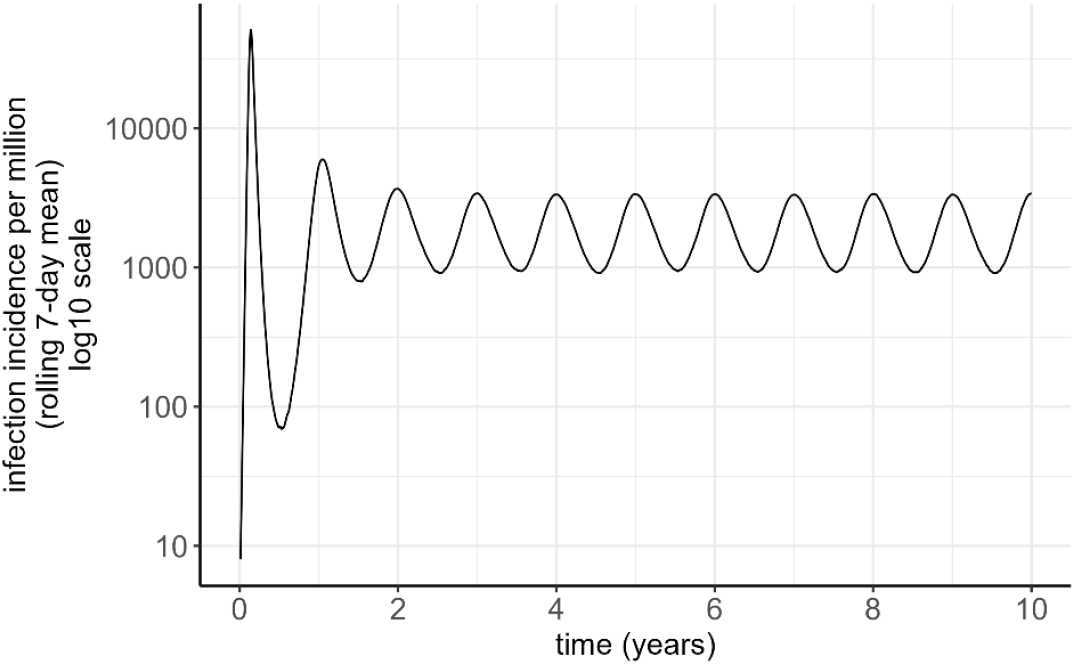
Infection incidence over time for a model simulation with no vaccination, and baseline parameters as specified in Table 1, illustrating infection endemicity reached within a 10-year burn-in period.

**Figure S3:**
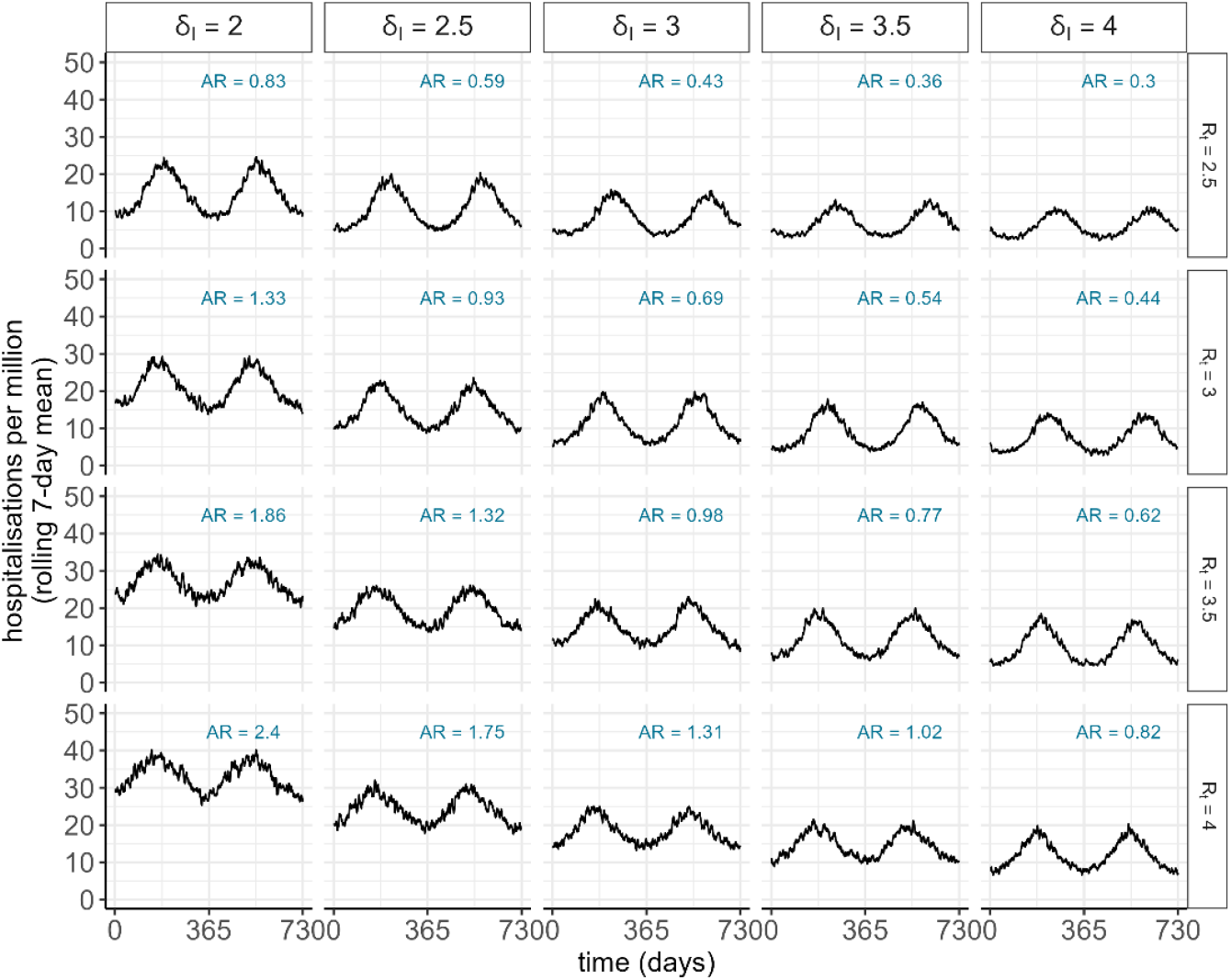
Model calibration without vaccination, across parameter sweeps varying the reproduction number *R*_*t*_, the amplitude of seasonal forcing *b*_1_, and the level of immune recognition following infection *δ*_*I*_. The attack rate (AR) is calculated as the total number of infections in a 12-month period divided by the total population.

**Figure S4:**
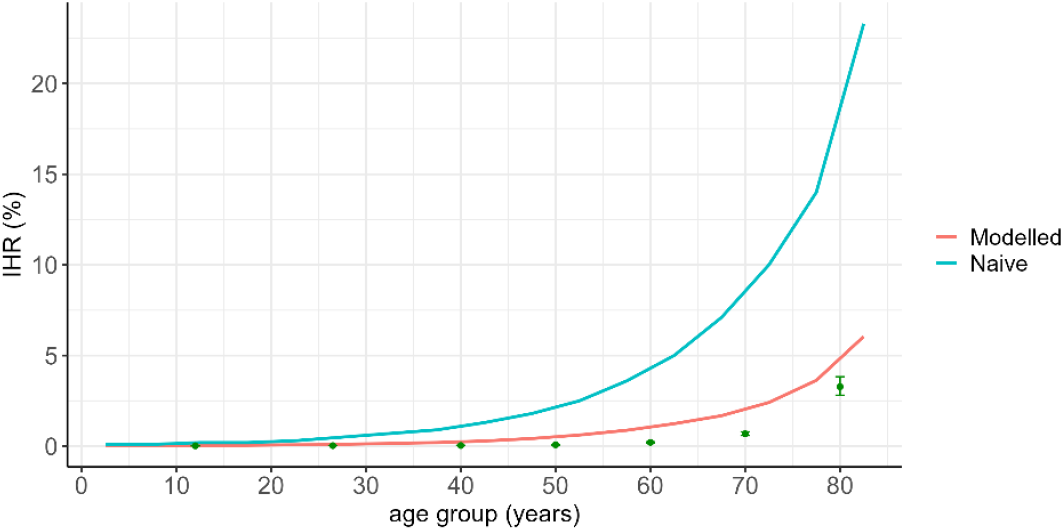
Infection hospitalisation ratio (IHR) (%) by age. The figure shows the model inputs (which represent the risk of hospitalisation with infection, for a “naïve” individual with no existing immunity), as the red line, and the modelled outputs as the green line, which represent the proportion of modelled infections that result in hospitalisation for a population with immunity. The blue dots depict recent estimates of the IHR for the UK, as published in the UK Health Security Agency “Winter Coronavirus (COVID-19) Infection Study: estimates of infection hospitalisation and fatality risk, 30 May 2024” (28).

**Figure S5.**
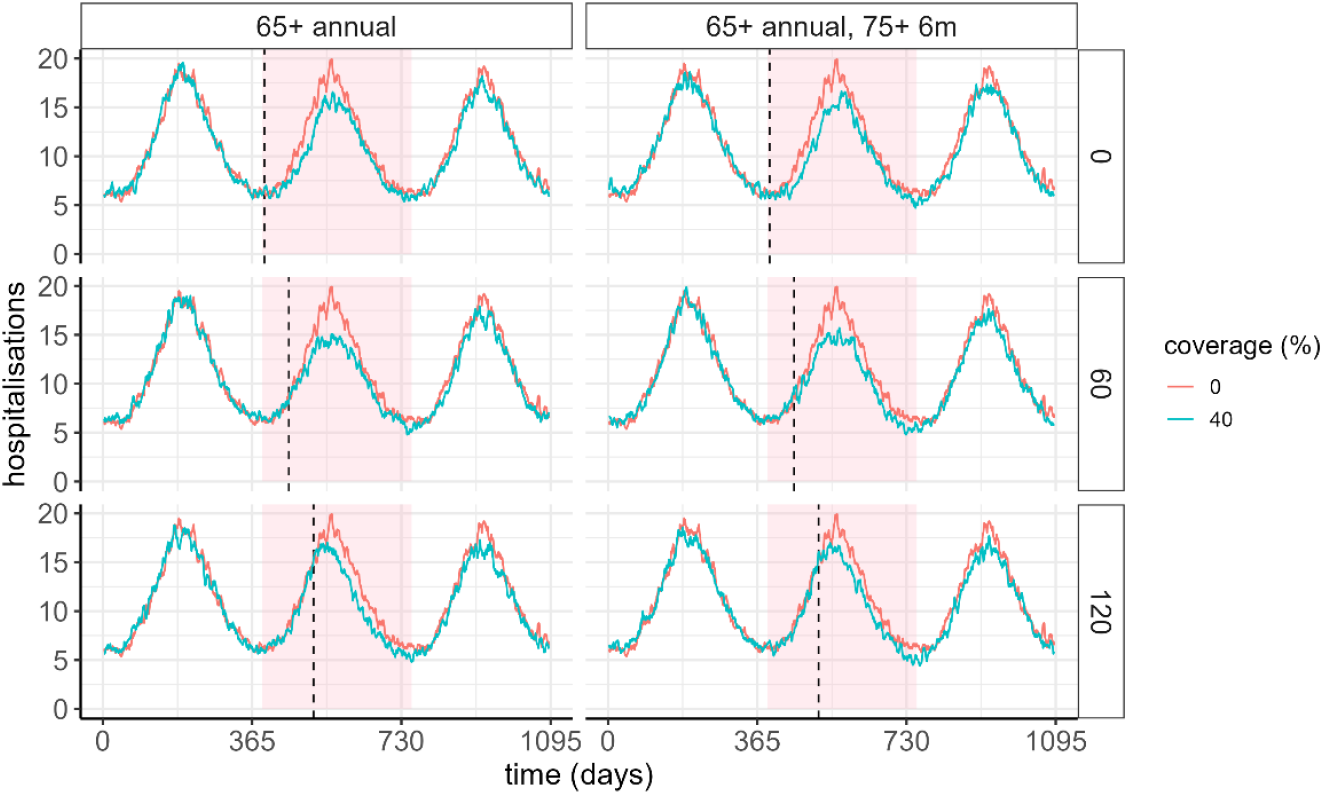
Vaccine impact shown as hospitalisations per day for a vaccine delivered at 40% coverage (blue line), relative to a scenario with no vaccination (red line), for a scenario with no additional immune escape. The vertical dashed line indicates the time of vaccination commencement, at either 0, 60, or 120 days after the trough in infections (rows). The pink shaded region denoted the time window over which results are aggregated in the main text. The columns represent different vaccine strategies: either a single dose delivered to the 65+ population, or a single dose delivered to the 65+ population with an additional dose to the 75+ population after 6 months.

**Figure S6.**
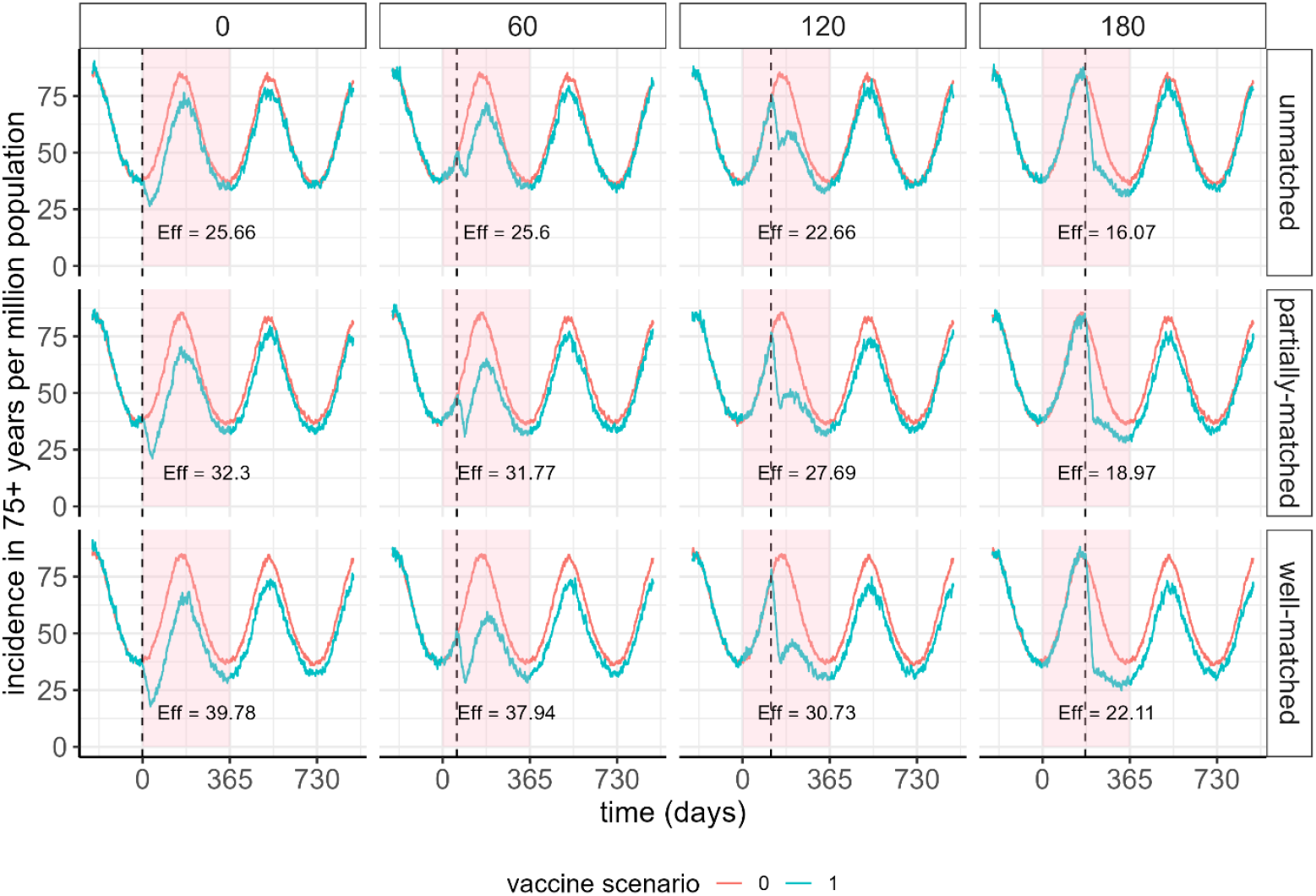
Modelled daily infections in the 75+ years age cohort, for a scenario with a vaccine delivered at high coverage in the 75+ years age cohort only (blue line), compared to a no-vaccination scenario. This is shown to demonstrate individual-level benefit of vaccination at different timing relative to the COVID-19 epidemic (columns), and for different levels of vaccine matching to the circulating SARS-CoV-2 strain (rows). The calculated vaccine effectiveness against hospitalisation over 12 months in the vaccinated cohort is annotated on each panel (%).

**Figure S7.**
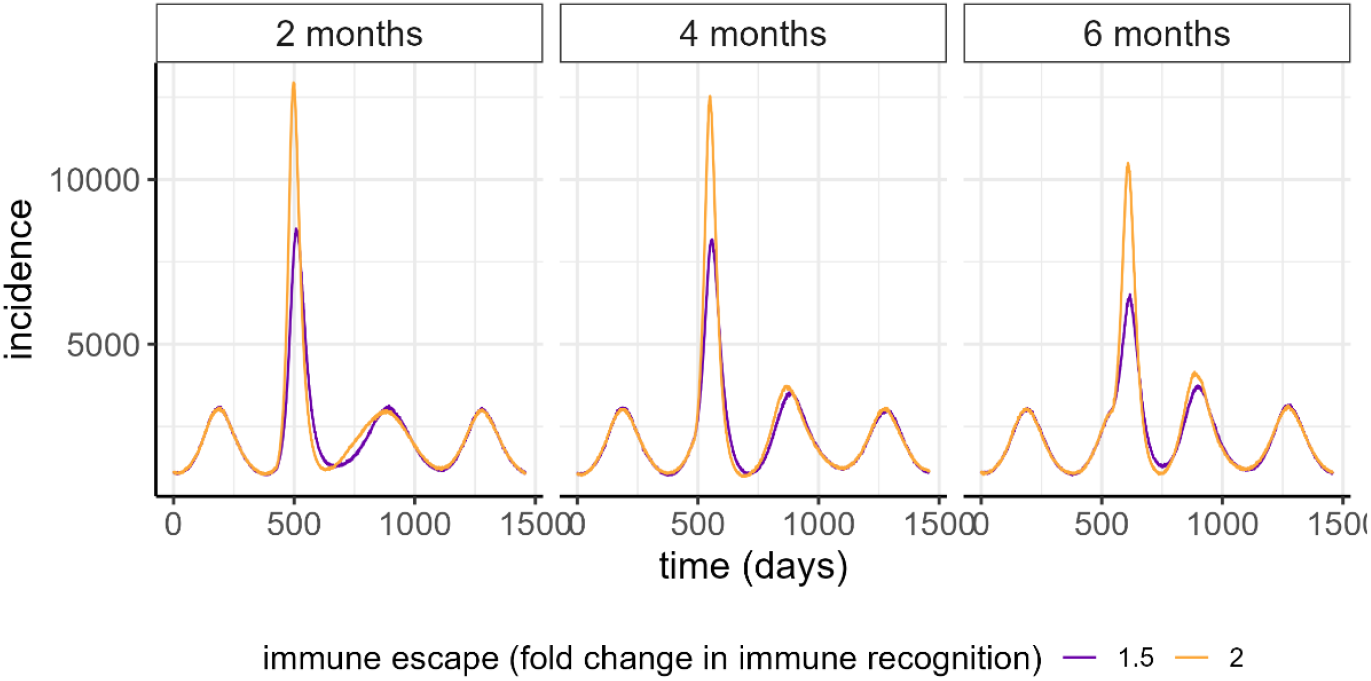
Simulated infections per day per million population, where an immune escape variant emerges either 2 months, 4 months, or 6 months following the seasonal trough (columns). Two levels of immune escape are simulated – a 1.5 (red) and 2 (blue) level fold drop in total immune recognition.

**Figure S8.**
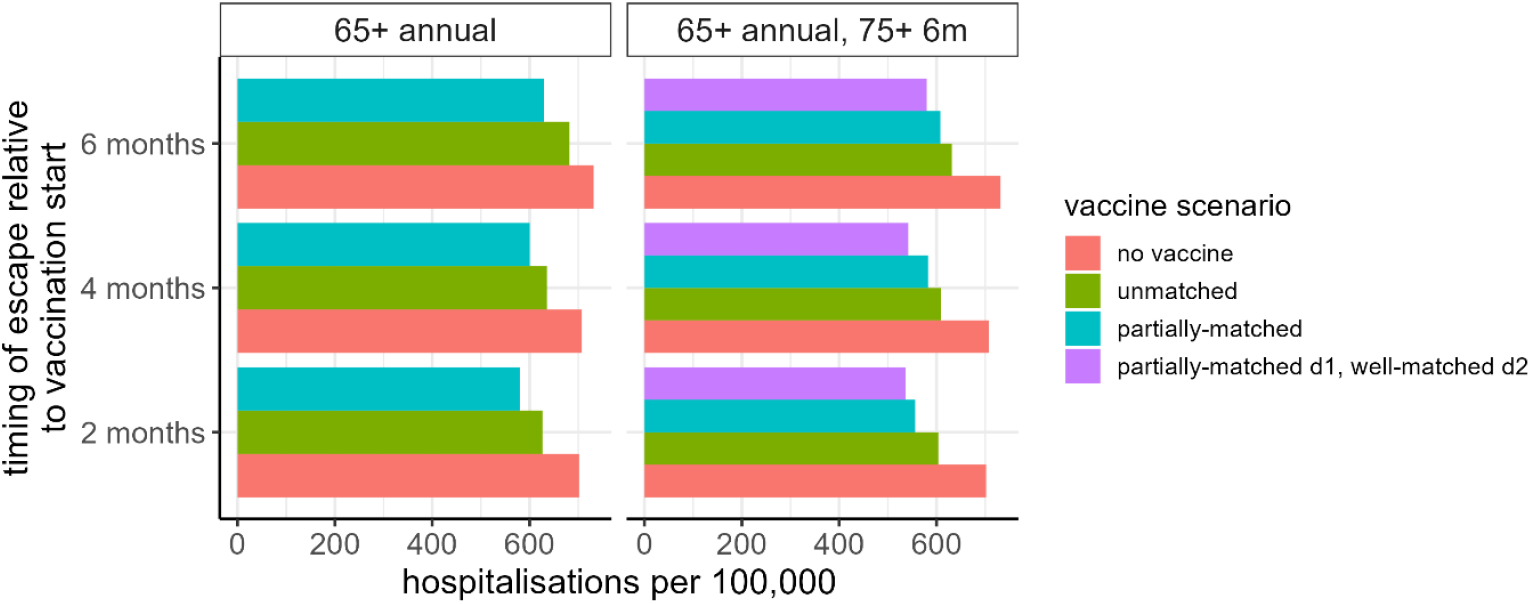
Total hospitalisations per 100,000 in the context of the emergence of a distinct variant strain either 2 months, 4 months, or 6 months following the seasonal trough (y-axis), for immune escape (a 1.5 level fold drop in immune recognition). Four vaccine scenarios are shown (coloured bars), for two vaccine delivery scenarios (columns): no vaccine (red); an unmatched vaccine (green), with *δ* = 1.5, a partially-matched vaccine, with *δ* = 2 (teal), and a partially matched first dose to the 65+ years population, with a second dose after 6 months delivered to the 75+ years population with a vaccine dose that is well-matched to the new variant strain (purple).

**Table S3:** Annual impact and averted events in terms of hospitalisations per 100,000 individuals, for routine vaccination scenarios. The modelled estimated baseline annual hospitalisations (i.e. in the no-vaccination group) was 417 per 100,000. Values represent the median across 20 model simulations.

| Vaccine matching | Coverage | Relative timing | Schedule: 65+ annual |  |  | Schedule: 65+ annual, 75+ 6m |  |  |
| --- | --- | --- | --- | --- | --- | --- | --- | --- |
|  |  |  | Hospitalisations per 100,000 | Averted per 100,000 | Proportion averted (%) | Hospitalisations per 100,000 | Averted per 100,000 | Proportion averted (%) |
| Unmatched | 20% | 0 days | 397 | 20 | 4.9 | 398 | 19 | 4.6 |
|  |  | 60 days | 404 | 13 | 3 | 401 | 16 | 3.8 |
|  |  | 120 days | 406 | 11 | 2.6 | 404 | 13 | 3.1 |
|  | 40% | 0 days | 385 | 32 | 7.7 | 383 | 34 | 8 |
|  |  | 60 days | 381 | 36 | 8.6 | 386 | 31 | 7.4 |
|  |  | 120 days | 393 | 24 | 5.7 | 393 | 24 | 5.7 |
|  | 60% | 0 days | 376 | 41 | 9.9 | 365 | 52 | 12.4 |
|  |  | 60 days | 371 | 46 | 10.9 | 369 | 48 | 11.6 |
|  |  | 120 days | 380 | 37 | 8.8 | 373 | 44 | 10.6 |
| Partially-matched | 20% | 0 days | 394 | 23 | 5.5 | 394 | 23 | 5.4 |
|  |  | 60 days | 395 | 22 | 5.2 | 389 | 28 | 6.7 |
|  |  | 120 days | 399 | 18 | 4.3 | 398 | 19 | 4.5 |
|  | 40% | 0 days | 376 | 41 | 9.9 | 365 | 52 | 12.4 |
|  |  | 60 days | 380 | 37 | 8.8 | 374 | 42 | 10.2 |
|  |  | 120 days | 383 | 34 | 8.3 | 381 | 36 | 8.7 |
|  | 60% | 0 days | 356 | 61 | 14.7 | 348 | 69 | 16.5 |
|  |  | 60 days | 363 | 54 | 12.9 | 351 | 66 | 15.7 |
|  |  | 120 days | 369 | 48 | 11.6 | 364 | 53 | 12.6 |
| Well-matched | 20% | 0 days | 390 | 27 | 6.5 | 381 | 36 | 8.6 |
|  |  | 60 days | 393 | 24 | 5.6 | 389 | 28 | 6.8 |
|  |  | 120 days | 397 | 20 | 4.9 | 400 | 17 | 4.2 |
|  | 40% | 0 days | 369 | 48 | 11.6 | 362 | 55 | 13.1 |
|  |  | 60 days | 367 | 50 | 12 | 365 | 52 | 12.5 |
|  |  | 120 days | 374 | 43 | 10.4 | 377 | 40 | 9.5 |
|  | 60% | 0 days | 347 | 70 | 16.8 | 335 | 82 | 19.7 |
|  |  | 60 days | 349 | 68 | 16.4 | 343 | 74 | 17.9 |
|  |  | 120 days | 355 | 62 | 14.9 | 361 | 56 | 13.3 |

**Table S4:** Impact of different vaccine schedules and products in the context of the emergence of a new variant strain, for different scenarios for timing of immune escape relative to vaccination. Impact is shown as annual total and averted hospitalisations per 100,000 individuals. Values represent the median across 20 model simulations.

| Timing of immune escape | Schedule | Vaccine matching | Baseline hospitalisations per 100,000 | Hospitalisations per 100,000 | Averted per 100,000 | Proportion averted (%) |
| --- | --- | --- | --- | --- | --- | --- |
| 2 months | 65+ annual | Unmatched | 702 | 627 | 75 | 10.7 |
|  |  | Partially-matched | 702 | 580 | 122 | 17.4 |
|  | 65+ annual, 75+ 6m | Unmatched | 702 | 603 | 98 | 14.1 |
|  |  | Partially-matched | 702 | 556 | 146 | 20.8 |
|  |  | Partially-matched d1, well-matched d2 | 702 | 536 | 166 | 23.6 |
| 4 months | 65+ annual | Unmatched | 707 | 636 | 72 | 10 |
|  |  | Partially-matched | 707 | 600 | 107 | 15.1 |
|  | 65+ annual, 75+ 6m | Unmatched | 707 | 609 | 98 | 13.9 |
|  |  | Partially-matched | 707 | 583 | 124 | 17.5 |
|  |  | Partially-matched d1, well-matched d2 | 707 | 542 | 165 | 23.3 |
| 6 months | 65+ annual | Unmatched | 732 | 682 | 50 | 6.8 |
|  |  | Partially-matched | 732 | 630 | 102 | 13.9 |
|  | 65+ annual, 75+ 6m | Unmatched | 732 | 631 | 100 | 13.8 |
|  |  | Partially-matched | 732 | 608 | 124 | 16.9 |
|  |  | Partially-matched d1, well-matched d2 | 732 | 580 | 152 | 20.8 |

## References

1. World Health Organization. WHO Director-General’s opening remarks at the media briefing – 5 May 2023 [Internet]. [cited 2024 Nov 22]. Available from: https://www.who.int/director-general/speeches/detail/who-director-general-s-opening-remarks-at-the-media-briefing5-may-2023

2. Andrews N, Tessier E, Stowe J, Gower C, Kirsebom F, Simmons R, et al. Duration of Protection against Mild and Severe Disease by Covid-19 Vaccines. N Engl J Med. 2022 Jan;386(4):340–50.

3. World Health Organization. COVID-19 deaths | WHO COVID-19 dashboard [Internet]. [cited 2024 Dec 2]. Available from: https://data.who.int/dashboards/covid19/cases

4. Australian Government Department of Health and Aged Care. COVID-19 reporting [Internet]. Australian Government Department of Health and Aged Care; 2024 [cited 2024 Dec 2]. Available from: https://www.health.gov.au/topics/covid-19/reporting

5. Australian Bureau of Statistics (ABS). Deaths due to COVID-19, influenza and RSV in Australia - 2022 - September 2024 | Australian Bureau of Statistics [Internet]. 2024 [cited 2024 Dec 3]. Available from: https://www.abs.gov.au/articles/deaths-due-covid-19-influenza-and-rsv-australia-2022-september-2024

6. Australian Government Department of Health and Aged Care. Australian Respiratory Surveillance Report 16 – 21 October 2024 to 17 November 2024 [Internet]. Australian Government Department of Health and Aged Care; 2024 [cited 2024 Dec 3]. Available from: https://www.health.gov.au/resources/publications/australian-respiratory-surveillance-report-16-21-october-2024-to-17-november-2024?language=en

7. Bergeri I, Whelan MG, Ware H, Subissi L, Nardone A, Lewis HC, et al. Global SARS-CoV-2 seroprevalence from January 2020 to April 2022: A systematic review and meta-analysis of standardized population-based studies. PLOS Med. 2022 Nov 10;19(11):e1004107.

8. Kirby Institute. Serosurveillance for SARS-CoV-2 infection to inform public health responses [Internet]. [cited 2024 Dec 3]. Available from: https://www.kirby.unsw.edu.au/research/projects/serosurveillance-sars-cov-2-infection-inform-public-health-responses

9. UK Health Security Agency. GOV.UK. 2024 [cited 2024 Dec 3]. COVID-19 vaccine quarterly surveillance reports (September 2021 to November 2024). Available from: https://www.gov.uk/government/publications/covid-19-vaccine-weekly-surveillance-reports

10. Mccartney, Kristine, Liu, Bette. Why older adults can continue to benefit from covid-19 boosters. BMJ. 2023 Jul 25;p1662.

11. World Health Organization. COVID-19 Vaccines Advice [Internet]. [cited 2024 Nov 22]. Available from: https://www.who.int/emergencies/diseases/novel-coronavirus-2019/covid-19-vaccines/advice

12. Australian Government Department of Health and Aged Care. COVID-19 vaccine advice and recommendations [Internet]. Australian Government Department of Health and Aged Care; 2024 [cited 2024 Nov 22]. Available from: https://www.health.gov.au/our-work/covid-19-vaccines/getting-your-vaccination

13. World Health Organization. Statement on the antigen composition of COVID-19 vaccines [Internet]. [cited 2025 Feb 7]. Available from: https://www.who.int/news/item/23-12-2024-statement-on-the-antigen-composition-of-covid-19-vaccines

14. UK Health Security Agency. GOV.UK. [cited 2024 Dec 5]. National flu and COVID-19 surveillance report: 28 November (week 48). Available from: https://www.gov.uk/government/statistics/national-flu-and-covid-19-surveillance-reports-2024-to-2025-season/national-flu-and-covid-19-surveillance-report-28-november-week-48

15. Australian Government Department of Health and Aged Care. COVID-19 vaccine rollout update – 8 November 2024 [Internet]. Australian Government Department of Health and Aged Care; 2024 [cited 2024 Dec 5]. Available from: https://www.health.gov.au/resources/publications/covid-19-vaccine-rollout-update-8-november-2024?language=en

16. Brooks-Pollock E, Danon L, Jombart T, Pellis L. Modelling that shaped the early COVID-19 pandemic response in the UK. Philos Trans R Soc B Biol Sci. 2021 May 31;376(1829):20210001.

17. Burch E, Khan SA, Stone J, Asgharzadeh A, Dawe J, Ward Z, et al. Early mathematical models of COVID-19 vaccination in high-income countries: a systematic review. Public Health. 2024 Nov 1;236:207–15.

18. Hogan AB, Doohan P, Wu SL, Mesa DO, Toor J, Watson OJ, et al. Estimating long-term vaccine effectiveness against SARS-CoV-2 variants: a model-based approach. Nat Commun. 2023 Jul 19;14(1):4325.

19. Hogan AB, Wu SL, Toor J, Mesa DO, Doohan P, Watson OJ, et al. Long-term vaccination strategies to mitigate the impact of SARS-CoV-2 transmission: A modelling study. PLOS Med. 2023 Nov 28;20(11):e1004195.

20. Khoury DS, Cromer D, Reynaldi A, Schlub TE, Wheatley AK, Juno JA, et al. Neutralizing antibody levels are highly predictive of immune protection from symptomatic SARS-CoV-2 infection. Nat Med. 2021 Jul;27(7):1205–11.

21. Lo Tartaro D, Paolini A, Mattioli M, Swatler J, Neroni A, Borella R, et al. Detailed characterization of SARS-CoV-2-specific T and B cells after infection or heterologous vaccination. Front Immunol. 2023 Feb 9;14.

22. GeurtsvanKessel CH, Geers D, Schmitz KS, Mykytyn AZ, Lamers MM, Bogers S, et al. Divergent SARS-CoV-2 Omicron–reactive T and B cell responses in COVID-19 vaccine recipients. Sci Immunol. 2022 Feb 3;7(69):eabo2202.

23. Wu, S, Watson, OJ, Winskill, P. Individual-Based Model of COVID Transmission [Internet]. [cited 2025 Mar 4]. Available from: https://mrc-ide.github.io/safir/

24. Canetti M, Barda N, Gilboa M, Indenbaum V, Asraf K, Gonen T, et al. Six-Month Follow-up after a Fourth BNT162b2 Vaccine Dose. N Engl J Med. 2022 Nov 30;387(22):2092–4.

25. Movsisyan M, Truzyan N, Kasparova I, Chopikyan A, Sawaqed R, Bedross A, et al. Tracking the evolution of anti-SARS-CoV-2 antibodies and long-term humoral immunity within 2 years after COVID-19 infection. Sci Rep. 2024 Jun 11;14(1):13417.

26. Srivastava K, Carreño JM, Gleason C, Monahan B, Singh G, Abbad A, et al. SARS-CoV-2-infection- and vaccine-induced antibody responses are long lasting with an initial waning phase followed by a stabilization phase. Immunity. 2024 Mar 12;57(3):587-599.e4.

27. Otto SP, MacPherson A, Colijn C. Endemic does not mean constant as SARS-CoV-2 continues to evolve. Evolution. 2024 Jun 1;78(6):1092–108.

28. UK Health Security Agency. GOV.UK. [cited 2024 Dec 12]. Winter Coronavirus (COVID-19) Infection Study: estimates of infection hospitalisation and fatality risk, 30 May 2024. Available from: https://www.gov.uk/government/statistics/winter-coronavirus-covid-19-infection-study-estimates-of-epidemiological-characteristics-england-and-scotland-2023-to-2024/winter-coronavirus-covid-19-infection-study-estimates-of-infection-hospitalisation-and-fatality-risk-30-may-2024

29. Kirwan PD, Foulkes S, Munro K, Sparkes D, Singh J, Henry A, et al. Protection of vaccine boosters and prior infection against mild/asymptomatic and moderate COVID-19 infection in the UK SIREN healthcare worker cohort: October 2023 to March 2024. J Infect. 2024 Nov 1;89(5):106293.

30. United Nations Population Division. World Population Prospects [Internet]. [cited 2024 Dec 12]. Available from: https://population.un.org/wpp/Download/Standard/MostUsed/

31. Mossong J, Hens N, Jit M, Beutels P, Auranen K, Mikolajczyk R, et al. Social contacts and mixing patterns relevant to the spread of infectious diseases. PLoS Med. 2008 Mar 25;5(3):e74.

32. Australian Government Department of Health and Aged Care. COVID-19 Vaccine Rollout Update 10 January 2025 [Internet]. 2025. Available from: https://www.health.gov.au/resources/collections/covid-19-vaccination-rollout-update

33. Australian Government Department of Health and Aged Care. COVID-19 Vaccine Rollout Update 10 January 2024 [Internet]. 2024. Available from: https://www.health.gov.au/resources/collections/covid-19-vaccination-rollout-update

34. Australian Government Department of Health and Aged Care. COVID-19 Vaccine Rollout Update 12 July 2024 [Internet]. 2024. Available from: https://www.health.gov.au/resources/publications/covid-19-vaccine-rollout-update-12-july-2024?language=en

35. Wang Y, Wei X, Liu Y, Li S, Pan W, Dai J, et al. Towards broad-spectrum protection: the development and challenges of combined respiratory virus vaccines. Front Cell Infect Microbiol. 2024;14:1412478.

36. Dobrzynski D, Branche AR, Falsey AR. Combination Seasonal Vaccines for Influenza, Respiratory Syncytial Virus, Severe Acute Respiratory Syndrome Coronavirus 2, and Other Pathogens. J Infect Dis. 2025 Feb 15;231(2):291–3.

37. Subissi L, Otieno JR, Worp N, Attar Cohen H, Oude Munnink BB, Abu-Raddad LJ, et al. An updated framework for SARS-CoV-2 variants reflects the unpredictability of viral evolution. Nat Med. 2024 Sep;30(9):2400–3.

38. Australian COVID-19 Serosurveillance Network. Seroprevalence of SARS-CoV-2-specific antibodies among Australian blood donors: Round 4 update [Internet]. 2023. Available from: https://www.kirby.unsw.edu.au/research/projects/serosurveillance-sars-cov-2-infection-inform-public-health-responses

39. GOV.UK [Internet]. [cited 2025 Mar 7]. Winter Coronavirus (COVID-19) Infection Study: estimates of epidemiological characteristics, 15 February 2024. Available from: https://www.gov.uk/government/statistics/winter-coronavirus-covid-19-infection-study-estimates-of-epidemiological-characteristics-england-and-scotland-2023-to-2024/winter-coronavirus-covid-19-infection-study-estimates-of-epidemiological-characteristics-15-february-2024

40. Queensland Government Department of Health. Acute respiratory infection surveillance reporting [Internet]. 2024 [cited 2024 Dec 11]. Available from: https://www.health.qld.gov.au/clinical-practice/guidelines-procedures/diseases-infection/surveillance/reports/flu

41. UK Health Security Agency. COVID-19 vaccine surveillance report: week 19 [Internet]. 2022. Available from: https://www.gov.uk/government/publications/covid-19-vaccine-weekly-surveillance-reports

42. Victorian Government Department of Health. Victorian COVID-19 surveillance report [Internet]. State Government of Victoria, Australia; [cited 2024 Dec 11]. Available from: https://www.health.vic.gov.au/infectious-diseases/victorian-covid-19-surveillancereport

43. Ankit Rohatgi. WebPlotDigitizer [Internet]. [cited 2024 Dec 19]. Available from: https://automeris.io

44. Hogan AB, Winskill P, Watson OJ, Walker PGT, Whittaker C, Baguelin M, et al. Within-country age-based prioritisation, global allocation, and public health impact of a vaccine against SARS-CoV-2: A mathematical modelling analysis. Vaccine. 2021 May 21;39(22):2995–3006.

45. Puhach O, Meyer B, Eckerle I. SARS-CoV-2 viral load and shedding kinetics. Nat Rev Microbiol. 2023 Mar;21(3):147–61.

